# Evaluating the Mental Health Impacts of the COVID-19 Pandemic in Urban South Africa: Perceived Risk of COVID-19 Infection and Childhood Trauma Predict Adult Depressive Symptoms

**DOI:** 10.1101/2020.06.13.20130120

**Authors:** Andrew Wooyoung Kim, Tawanda Nyengerai, Emily Mendenhall

## Abstract

South Africa’s national lockdown introduced serious threats to public mental health in a society where one in three individuals develop a psychiatric disorder during their life. We aimed to evaluate the mental health impacts of the COVID-19 pandemic using a mixed methods design. This longitudinal study drew from a preexisting sample of 957 adults living in Soweto, a major township near Johannesburg. Psychological assessments were administered across two waves: between August 2019-March 2020 and during the first six weeks of the lockdown (late March-early May 2020). Interviews on COVID-19 experiences were administered in the second wave. Multiple regression models examined relationships between perceived COVID-19 risk and depression. Full data on perceived COVID-19 risk, depression, and covariates were available in 221 adults. 14.5% of adults were at risk for depression. Higher perceived COVID-19 risk predicted greater depressive symptoms (*p* < 0.001) particularly among adults with histories of childhood trauma, though this effect was marginally significant (*p* = 0.062). Adults were two times more likely to experience significant depressive symptoms for every one unit increase in perceived COVID-19 risk (*p* = 0.016; 95% CI [1.14, 3.49]). Qualitative data identified potent experiences of anxiety, financial insecurity, fear of infection, and rumination. Higher perceived risk of COVID-19 infection is associated with greater depressive symptoms among adults with histories of childhood trauma during the first six weeks of quarantine. High rates of severe mental illness and low availability of mental healthcare amidst COVID-19 emphasize the need for immediate and accessible psychological resources in South Africa.

## INTRODUCTION

The South African response to coronavirus was swift and assertive in testing, tracing, and quarantining those infected with COVID-19. Despite the rapid and effective public health response^1^ the economic and social ramifications of quarantine have disproportionately afflicted those already socioeconomically disadvantaged in a society defined by its racial and economic inequity. Furthermore, harsh government sanctions to adhere to COVID-19 mitigation plans, including militarization, demolitions of informal settlements, and widespread police brutality have exacerbated already vulnerable communities who are unable to properly quarantine. These measures bring focus to the existing disproportionate inequalities in common mental disorders that are amplified by social distancing, strict quarantine measures, and the countrywide lockdown.

The South African government imposed a strict and militarized “national lockdown” policy on March 26, 2020 that prohibited citizens from leaving quarantine except for food, medicine, and essential labor. Worldwide, numerous aspects of life under forced confinement, including the limited physical mobility, emotional distress, and for some, extreme threats to survival, have substantially increased risk for mental distress and illness^2^. Studies on the mental health consequences of quarantine worldwide have reported marked increases in risk for depression, anxiety, post-traumatic stress disorder, and suicide^3^. For millions of South Africans, vulnerability to COVID-19 infection is amplified by other pre-existing adversities, such as hunger and violence, an overburdened healthcare system, a high prevalence of chronic and infectious disease, and alarming rates of poverty (55.5%) and unemployment (29%)^4^.

Based on evidence from recent pandemics (e.g. SARS, MERS), poorer mental health status before quarantine is a major risk factor for exacerbating the severity and duration of psychiatric morbidity after quarantine^5,6^. Recent estimates show that the prevalence, incidence, and burden of mental illness in South Africa are relatively high compared to other countries: one in three (30.3%) South Africans will be diagnosed with a mental illness, a quarter of all cases (25%) are considered severe, and nearly half of citizens (47.5%) are at risk of developing a psychiatric disorder in their lifetime^7^. Despite these conditions, mental healthcare usage and access in South Africa is severely limited, with only 27% of patients with severe mental illnesses receiving treatment, 16% of citizens enrolled in medical aid, and only 0.31 psychiatrist per 100,00 uninsured population^8^. These severe barriers to care amidst high rates of mental illness suggest that mental healthcare must be a priority for thinking about how people experience and respond to quarantine. The limited capacity of government public health initiatives and lack of research on disease burdens of COVID-19 highlight the urgent need for additional screening, treatment, and research efforts nationwide.

To address these gaps, this study investigates the mental health impacts of the COVID-19 pandemic among adults residing in Soweto, a major township southwest of Johannesburg, during the South African lockdown of 2020. Specifically, our analyses examine the relationships between perceived risk of COVID-19 infection and depressive symptoms and perceptions of COVID-19 and mental health. During the first six weeks of lockdown, we conducted follow-up, mixed-method interviews with adults enrolled in an existing epidemiological surveillance study to assess experiences of COVID-19 and adult depressive symptoms. To date, no study has empirically evaluated the mental health effects of COVID-19 experiences in South Africa.

## METHODS

### Study setting

This research was nested within the Developmental Pathways for Health Research Unit, which is part of the University of the Witwatersrand and located at Chris Hani Baragwanath Academic Hospital located in Soweto, South Africa. Soweto is a low-income neighborhood within the expansive land-locked city of Johannesburg, South Africa, famous for its incorporation of six townships. Today more than one million people reside in Soweto and most are black South Africans, representing various ethnic identities (e.g. Zulu, Xhosa, Sotho, etc.). Soweto is diverse economically, including middle-class neighborhoods, working class communities, and informal settlements. Residents report an elevated affliction of infectious conditions like HIV and TB and non-communicable diseases, such as hypertension, type 2 diabetes, and depression. The prevalence of multimorbidity is high, which is compounded by costly health care services in the private sector and systemic barriers in the public sector.

### Sample characteristics

All research participants were residents of Soweto and enrolled in a study preexisting the coronavirus pandemic. The first study was an epidemiological surveillance study of comorbidities, including mental (e.g. depression, anxiety), infectious (e.g. TB, HIV), and cardiometabolic diseases. All participants were 25 years or older and represented a wide range of ages and socioeconomic status, though a majority of our sample were women (Table 1). Participants during the first wave of data collection were interviewed in their homes and provided informed consent. Participants were recruited based on a simple random sample of geographic coordinates within the boundaries of Soweto (n=957). The University of the Witwatersrand Human Research Ethics Council reviewed and approved the study.

**Table 1.**
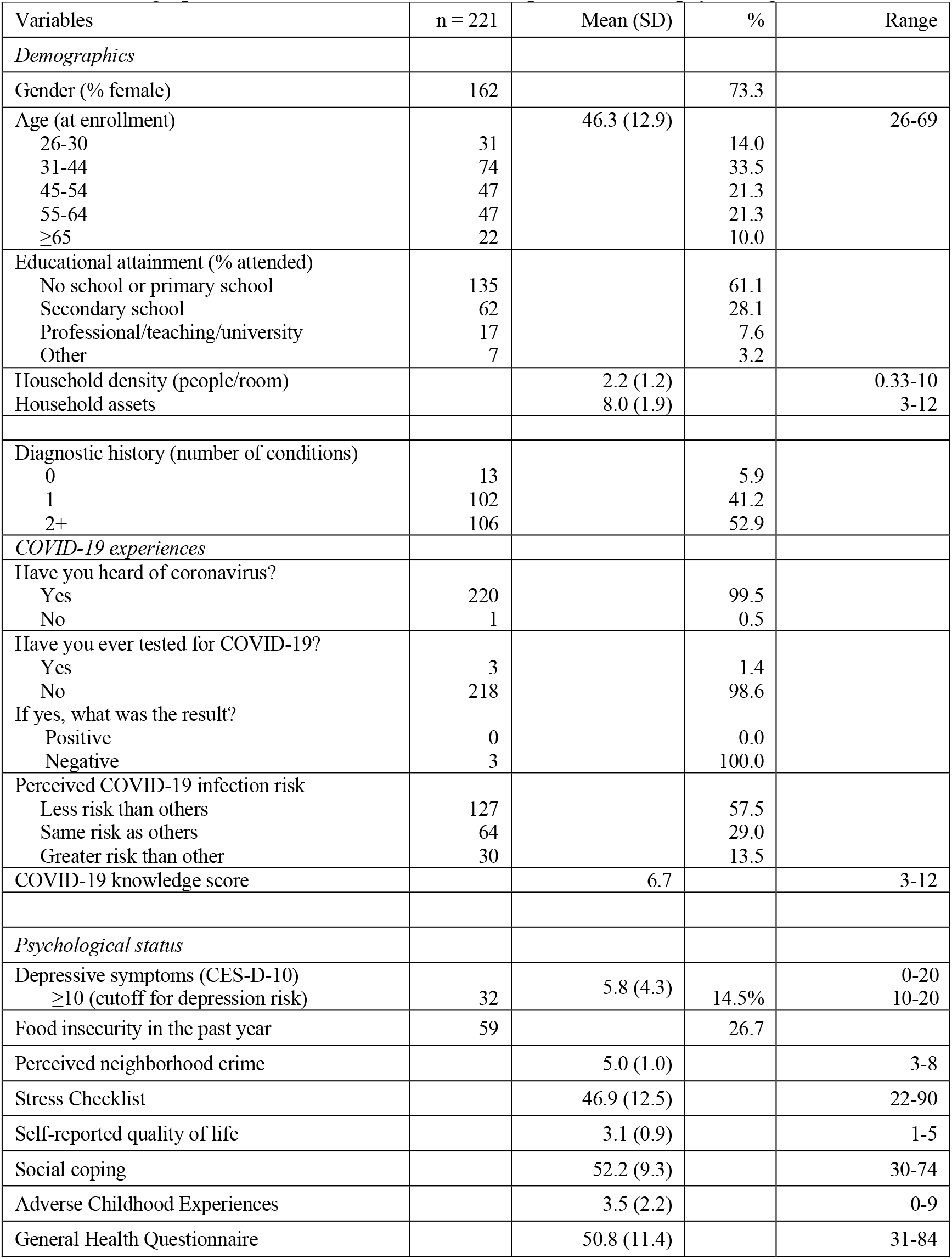
Demographic characteristics, COVID-19 experiences, and psychological status

| Variables | n = 221 | Mean (SD) | % | Range |
| --- | --- | --- | --- | --- |
| *Demographics* |  |  |  |  |
| Gender (% female) | 162 |  | 73.3 |  |
| Age (at enrollment) |  | 46.3 (12.9) |  | 26-69 |
| 26-30 | 31 |  | 14.0 |  |
| 31-44 | 74 |  | 33.5 |  |
| 45-54 | 47 |  | 21.3 |  |
| 55-64 | 47 |  | 21.3 |  |
| ≥65 | 22 |  | 10.0 |  |
| Educational attainment (% attended) |  |  |  |  |
| No school or primary school | 135 |  | 61.1 |  |
| Secondary school | 62 |  | 28.1 |  |
| Professional/teaching/university | 17 |  | 7.6 |  |
| Other | 7 |  | 3.2 |  |
| Household density (people/room) |  | 2.2 (1.2) |  | 0.33-10 |
| Household assets |  | 8.0 (1.9) |  | 3-12 |
| Diagnostic history (number of conditions) |  |  |  |  |
| 0 | 13 |  | 5.9 |  |
| 1 | 102 |  | 41.2 |  |
| 2+ | 106 |  | 52.9 |  |
| *COVID-19 experiences* |  |  |  |  |
| Have you heard of coronavirus? |  |  |  |  |
| Yes | 220 |  | 99.5 |  |
| No | 1 |  | 0.5 |  |
| Have you ever tested for COVID-19? |  |  |  |  |
| Yes | 3 |  | 1.4 |  |
| No | 218 |  | 98.6 |  |
| If yes, what was the result? |  |  |  |  |
| Positive | 0 |  | 0.0 |  |
| Negative | 3 |  | 100.0 |  |
| Perceived COVID-19 infection risk |  |  |  |  |
| Less risk than others | 127 |  | 57.5 |  |
| Same risk as others | 64 |  | 29.0 |  |
| Greater risk than other | 30 |  | 13.5 |  |
| COVID-19 knowledge score |  | 6.7 |  | 3-12 |
| *Psychological status* |  |  |  |  |
| Depressive symptoms (CES-D-10) |  | 5.8 (4.3) |  | 0-20 |
| ≥10 (cutoff for depression risk) | 32 |  | 14.5% | 10-20 |
| Food insecurity in the past year | 59 |  | 26.7 |  |
| Perceived neighborhood crime |  | 5.0 (1.0) |  | 3-8 |
| Stress Checklist |  | 46.9 (12.5) |  | 22-90 |
| Self-reported quality of life |  | 3.1 (0.9) |  | 1-5 |
| Social coping |  | 52.2 (9.3) |  | 30-74 |
| Adverse Childhood Experiences |  | 3.5 (2.2) |  | 0-9 |
| General Health Questionnaire |  | 50.8 (11.4) |  | 31-84 |

### Demographic, Health, and Socioeconomic Variables

Wave 1 involved an extensive demographic survey which queried age, gender, household conditions, education, and disease history. Household socioeconomic status (SES) was assessed using an asset index which scored participants according to the number of household physical assets they possessed out of 12 items, which was designed based on standard measures from the Demographic and Health Surveys (https://dhsprogram.com/). Food insecurity was assessed by asking whether household members experienced hunger in the past 12 months. Safety was assessed by summing two questions about how safe participants felt walking around their neighborhood during the day and night using a 4-point likert scale (very unsafe to very safe).

### COVID-19 experiences survey

Wave 2 involved a mixed-methods survey that was created during the weeks prior to the national lockdown and administered telephonically. Our COVID-19 experiences survey assessed awareness of COVID-19, COVID-19 infection status, and testing history. We assessed perceptions of COVID-19 prevention strategies, which asked whether a series of social and health behavior practices was understood to prevent and decrease risk of infection (e.g. can you get infected by being around people who cough/sneeze, sharing meals; can you prevent transmission by wearing a face mask, social distancing, etc.) to which participants responded “yes” or “no.” We summed the number of correct answers to create a composite measure of “COVID-19 knowledge” (12 items). The internal consistency of the knowledge measure was 0.78. Perceived risk of COVID-19 infection was assessed by asking “Do you think you have the same risk as others?” Participants indicated whether they had less, the same, or more risk than others.

### Psychological screeners

#### Wave 1

The General Health Questionnaire (GHQ-28) is a psychological screener that provides a measure of psychiatric risk based on four 7-item scales: somatic symptoms, anxiety and insomnia, social dysfunction, and severe depression. The survey assesses changes in mood, feelings, and behaviors in the past four weeks. Individuals evaluate their occurrence on a 4-point likert scale. Seven questions are reverse scored and transformed before all responses summed. The internal consistency was 0.86.

We evaluated stress and coping through three scales; two were created based on previous ethnographic work^9,10^. First, the Soweto Coping Scale (SCS) was a 14-item measure that assessed various coping behaviors, ranging from individual psychological practices, family and peer support, and religious activities^11^. She SCS had an internal consistency of 0.71. Second, Soweto Stress Scale (SSS)^12^ was a 21-item measure that assessed the severity of stressful experiences due to personal concerns, interpersonal conflict, family strife, economic deprivation, community safety, and violence. The internal consistency was 0.80. Third, the Adverse Childhood Experiences Study (ACES) questionnaire retrospectively assesses experiences of abuse, neglect, and household dysfunction during childhood. Participants provide yes/no responses to queries about ten distinct adverse events in their upbringing. Finally, we used the question, “how would you rate your quality of life?” as a generalized measure of well-being. At the end of each interview, we offered resources for free telephone-based psychological counseling at a major mental health NGO in Johannesburg. Research assistants were encouraged to use these resources weekly due to potential psychological burden of data collection.

#### Wave 2

The 10-item Center for Epidemiologic Studies Depression (CES-D) Scale assesses major symptoms of depression: depressed mood, changes in appetite and sleep, low energy, feelings of hopelessness, low self-esteem, and loneliness. Respondents considered the presence and duration of each item/symptom over the past week and rated each along a 4-point scale from 0 (rarely or never) to 3 (most or all of the time). Possible scores range from 0 to 30: a score of 10 and above indicates the presence of significant depressive symptoms. We found the CES-D had an internal consistency of 0.78.

### Statistical analyses

All analyses were conducted using version 15.1 of Stata (Stata Corporation, College Station, TX). All variables were examined for normal distribution and outliers. Bivariate analyses were conducted between CES-D scores, perceived COVID risk, and covariates. With the exception of known psychological, household, and social factors that may confound the relationship between perceived COVID-19 risk and depression, only those that were statistically significant at the 0.1 level during bivariate analyses were included final models. The following variables were included in the final model: gender, age, SES, household density (inhabitants/rooms), psychiatric risk (GHQ-28), childhood trauma, coping ability, and self-reported quality of life. All covariates were assessed during the first wave of data collection. General psychosocial stress was evaluated for possible inclusion but was removed because of high covariance with existing covariates. Food insecurity, safety, and chronic illness status were considered but removed because their associations were not significant at the 0.1 level. Multiple ordinary least squares (OLS) regressions were conducted to examine the relationship between perceived COVID-19 and depressive symptoms.

## RESULTS

Complete data on perceived COVID-19 risk, depression, and covariates were available for 221 adults (Table 1). Participants included in the analytical sample were similar to those excluded (n = 113) with respect to depression scores (CES-D-10), gender, age, assets, density, coping scores, quality of life, psychiatric risk (GHQ-28), and adverse childhood experiences (*p* > 0.05). Perceived COVID-19 risk was significantly different from those excluded from the sample (*p* < 0.05). Adults in the analytical sample exhibited higher perceived risk of COVID-19 infection. Using the CES-D-10 cutoff score of 10, 14.5% of adults in our sample were at risk for major depressive disorder (MDD).

Table 2 shows the most common qualitative responses to the open-ended question, “How do you think COVID-19 affects the mind^*^?” We found that most people said “no” (74%), COVID-19 does not affect the mind. Twenty percent indicated that COVID-19 causes what we coded as “anxiety” reflected by “there is so much fear since it started, you hear the increase in numbers of infected people everyday, you have no idea when it will come near you.” Other responses were “worries when it will stop,” “fear and panic,” and “fear of the future.” Some indicated that the infection itself would cause mental distress (16%), such as “the virus is a scary thought” and “I’m very afraid, being HIV+ I am very afraid of contracting corona because it might kill me.” Next, many people described “thinking too much” (10%), such as “coronavirus affects my mind because it is something that we are always thinking about.” The “lockdown” itself affected some (9%), such as “there are many restrictions” and “most people around the area don’t seem to be complying to the regulations.” Some spoke of financial stress (8%), such as “we cannot make a decent money under lockdown restrictions” and “no job brings in an income, food is now scarce.” Other common responses addressed stress like “it has added more stress since there were a lot of issues to deal with, for example the lack food for kids going to school.” Other challenges reported independently and overlapping with these primary responses include sadness that you cannot socialize, family stress and death – including not attending a loved one’s funeral, coping with the isolation of quarantine, and the chronic uncertainty of the days to come.

**Table 2.** Assessment of knowledge on COVID-19 transmission and prevention

| Question | # of correct answers | % (n=221) |
| --- | --- | --- |
| *Do you get COVID-19 from...* |  |  |
| Touching others | 114 | 52 |
| Being around others who cough | 114 | 52 |
| Being around others who sneeze | 186 | 84 |
| Sharing meals | 10 | 4.5 |
| *What will help prevent COVID-19 transmission?* |  |  |
| Hand washing only with water (Reverse coded) | 218 | 98.6 |
| Hand washing with water and soap | 209 | 94.6 |
| Covering your mouth when coughing/sneezing | 107 | 48.4 |
| Staying home | 147 | 66.5 |
| Drinking water (Reverse coded) | 186 | 84.2 |
| Wearing a face mask | 52 | 23.5 |
| Disinfecting surfaces | 59 | 26.7 |
| Social distancing | 75 | 33.9 |

Table 3 presents the results of the OLS regression analyses of demographic, household, psychological, and environmental factors that predict depressive symptoms. The unadjusted model (Model l) predicting CES-D scores on perceived COVID-19 risk displays a positive significant relationship (β = 1.6, *p* = <0.001, 95% CI [0.79, 2.34]). This relationship between COVID-19 risk and depression remains highly significant after adjusting for gender, age, SES, and household density (*p* = <0.001) (Models 2-5). Models 6 through 10 examine the potential confounding effects of psychosocial experiences and behaviors in shaping the relationship between perceived COVID-19 risk and depressive symptoms. The effect of COVID-19 risk slightly weakens (β = 1.31, *p* = 0.001, 95% CI [0.53, 2.08]) after adjusting for past psychiatric risk assessed through the GHQ-28, which is positively and significantly associated with CES-D scores (*p* = <0.001) (Model 6). Adding childhood trauma into the model (Model 7) very modestly weakens the perceived COVID-19 risk coefficient (β = 1.30, *p* = 0.001, 95% CI [0.54, 2.07]). Social coping behavior is inversely and insignificantly related to CES-D scores (*p* = 0.872) (Model 8). Model 9 and 10 shows that self-reported quality of life is negatively and significantly related to adult depression symptoms after the lockdown (*p* = 0.022), while COVID-19 knowledge positive and significantly predicts CES-D scores (*p* = 0.003) and also strengthens the COVID-19 risk coefficient (β = 1.48, *p* = <0.001, 95% CI [0.73, 2.23]).

**Table 3.** Relative Frequency Distribution of Qualitative Data

| Code | Frequency | % | Quote |
| --- | --- | --- | --- |
| No, COVID does not affect my mental health | 164 | 74.2 | No, I only think about it if I hear other people talking about it. I don't focus my mind on it. |
| Anxiety | 44 | 19.9 | Yes, now that my mother is not feeling well, I'm just worried as I don't know the signs and symptoms. |
| Infection | 33 | 14.9 | Yes, I'm very afraid, being HIV+ I am very afraid of contracting corona because it might kill me. |
| Thinking too much | 21 | 9.5 | Yes, I do think that coronavirus affects my mind because it is something that we are always thinking about. We are always scared, especially when we have to go out of the house. |
| Lockdown | 20 | 9.0 | Yes, we cannot make a decent living under these lockdown conditions. |
| Financial problems | 18 | 8.1 | Yes, I'm worried that I'm not working, my fiancé left, I'm in financial burden. I have a child who still needs provision. |
| Stress | 15 | 6.9 | Yes, there is a lot of fear and uncertainty regarding food security, education, and our general wellbeing. |
| Can't socialize | 13 | 5.9 | Yes, I'm feeling lonely. |
| Coping | 11 | 5.0 | Yes, but I have accepted that it's life. |
| Family | 11 | 5.0 | Yes, but I'm with my family so we are there for one another. |
| Death | 10 | 4.5 | Yes, everyone speaks about it everywhere. Now that the Government has no cure for it and it causes death. |
| Other | 39 | 17.2 | Yes, since I am a religious person, now I cannot be in fellowship with other people. |

**Table 4.** Multiple regression models of perceived COVID-19 risk predicting adult depression

|  | Model 1 | Model 2 | Model 3 | Model 4 | Model 5 | Model 6 | Model 7 | Model 8 | Model 9 | Model 10 |
| --- | --- | --- | --- | --- | --- | --- | --- | --- | --- | --- |
| Perceived COVID-19 risk | 1.6 ± 0.39*** | 1.6 ± 0.40*** | 1.5 ± 0.4*** | 1.5 ± 0.4*** | 1.6 ± 0.4*** | 1.3 ± 0.4*** | 1.3 ± 0.4*** | 1.3 ± 0.4*** | 1.4 ± 0.4*** | 1.5 ± 0.4*** |
| Gender (female) |  | 0.2 ± 0.6 | 0.2 ± 0.6 | 0.2 ± 0.6 | 0.3 ± 0.7 | 0.1 ± 0.6 | 0.1 ± 0.6 | 0.2 ± 0.6 | 0.3 ± 0.6 | 0.1 ± 0.6 |
| Maternal age |  |  | 0.003 ± 0.2 | 0.002 ± 0.2 | -0.0001 ± 0.02 | 0.003 ± 0.02 | 0.003 ± 0.02 | 0.008 ± 0.02 | 0.003 ± 0.02 | 0.01 ± 0.02 |
| Asset |  |  |  | -0.05 ± 0.2 | -0.05 ± 0.2 | -0.05 ± 0.2 | -0.05 ± 0.2 | 0.001 ± 0.2 | 0.009 ± 0.2 | -0.02 ± 0.1 |
| Density |  |  |  |  | -0.2 ± 0.2 | -0.2 ± 0.2 | -0.2 ± 0.2 | -0.2 ± 0.2 | -0.2 ± 0.2 | -0.2 ± 0.2 |
| Psychiatric risk |  |  |  |  |  | 0.1 ± 0.02*** | 0.1 ± 0.03*** | 0.08 ± 0.03*** | 0.07 ± 0.03* | 0.08 ± 0.03** |
| Childhood trauma |  |  |  |  |  |  | 0.3 ± 0.1* | 0.3 ± 0.1* | 0.3 ± 0.1* | 0.2 ± 0.1 |
| Coping |  |  |  |  |  |  |  | -0.004 ± 0.03 | 0.004 ± 0.03 | 0.008 ± 0.03 |
| Quality of life |  |  |  |  |  |  |  |  | -0.8 ± 0.3* | -0.8 ± 0.3* |
| COVID-19 knowledge |  |  |  |  |  |  |  |  |  | 0.4 ± 0.1** |
| Intercept | 4.9 ± 0.36*** | 4.8 ± 0.58*** | 4.6 ± 1.1*** | 5.1 ± 1.8*** | 5.6 ± 1.9*** | 0.5 ± 2.2 | -0.2 ± 2.2 | 0.02 ± 2.7 | 3.0 ± 3.0 | -0.5 ± 3.1 |
| Model R<sup>2</sup> | 0.0673 | 0.0678 | 0.0679 | 0.0683 | 0.0711 | 0.1383 | 0.1554 | 0.1555 | 0.1799 | 0.2135 |

† $p < 0.1$ \* $p < 0.05$ \*\* $p < 0.01$ \*\*\* $p < 0.001$

|  | Model 11 (with interaction) |
| --- | --- |
| Perceived COVID-19 risk | 0.3 ± 0.7 |
| Childhood trauma | -0.01 ± 0.2 |
| Perceived COVID-19 risk x Childhood trauma | 0.3 ± 0.2† |
| Gender (female) | 0.09 ± 0.6 |
| Maternal age | 0.008 ± 0.02 |
| Asset | 0.002 ± 0.1 |
| Density | -0.2 ± 0.2 |
| Psychiatric risk | 0.08 ± 0.03** |
| Coping | 0.01 ± 0.03 |
| Quality of life | -0.8 ± 0.3* |
| COVID knowledge | 0.4 ± 0.1** |
| Intercept | -0.03 ± 3.2 |
| Model R<sup>2</sup> | 0.2265 |

† $p < 0.1$ \* $p < 0.05$ \*\* $p < 0.01$ \*\*\* $p < 0.001$

Figure 1 illustrates the positive relationship between perceived risk of COVID-19 infection and depression scores in our fully adjusted model: greater perceived risk of COVID-19 infection corresponds with greater depressive symptoms in our sample. Elevated psychiatric risk, childhood trauma, and a greater degree of “COVID-19 knowledge” were positive and significant predictors of worse depressive symptoms, while higher quality of life was inversely and significantly related. The fully adjusted model (Model 10) accounts for 21% of the variance in depressive symptoms. Additionally, a logistic regression of depression risk (using CES-D ≥10 as a cutoff for significant depressive symptoms) on perceived COVID-19 risk with identical covariates estimated that the odds of the presence of significant depressive symptoms is 1.99 (*p* = 0.016; 95% CI [1.14, 3.49]) for every one unit increase in perceived COVID-19 risk.

**Figure 1.**
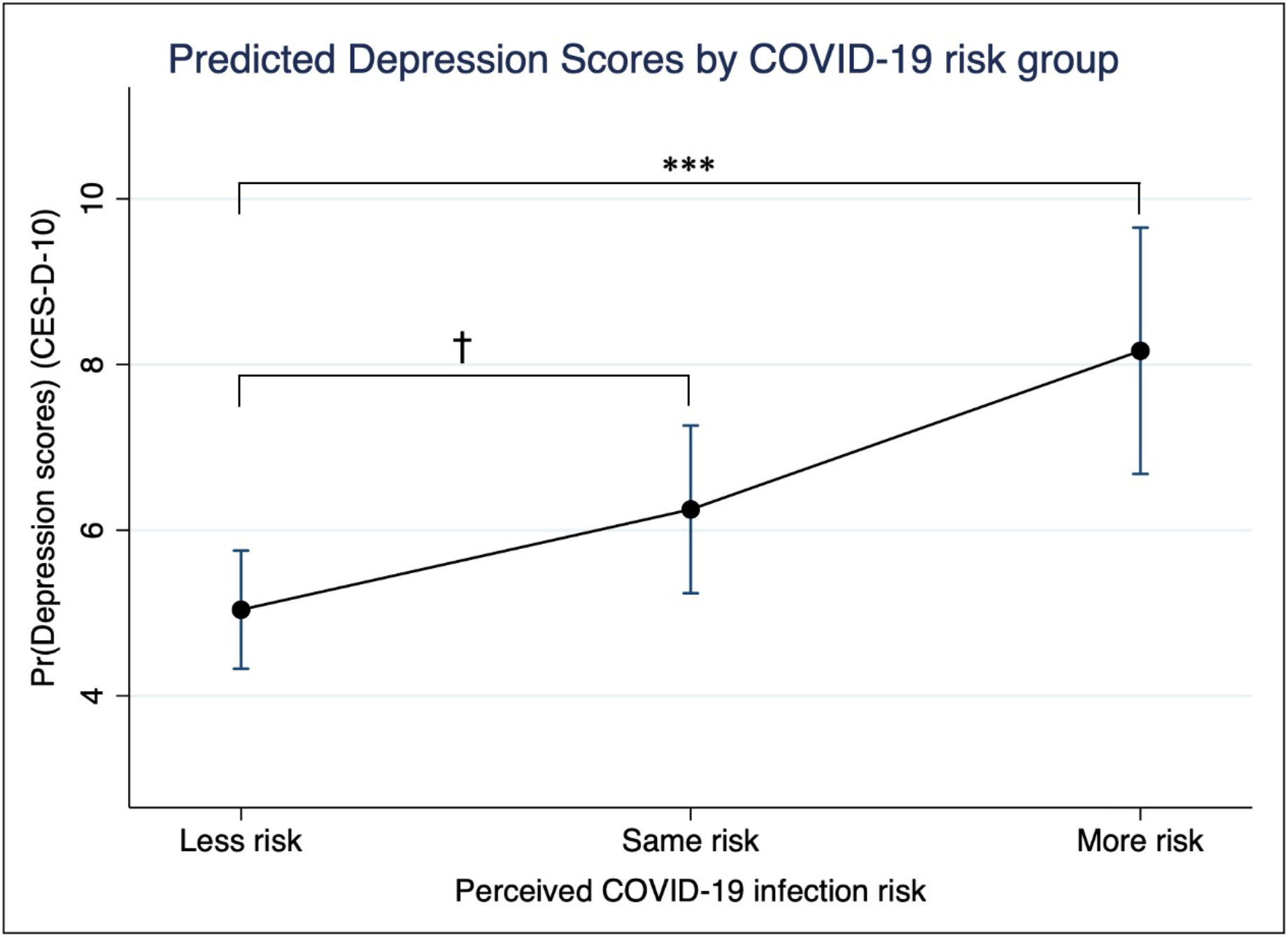
Predicted depression scores by perceived COVID-19 risk group. *Note:* Greater perceived risk of COVID-19 infection corresponds with greater depression symptomatology in adults living in Soweto. The effect of being in the “More risk” group is highly significant (*p* = <0.001) relative to being at “Less risk”, while the effect of perceiving that one is at the “Same risk” of COVID-19 infection relative to other individuals living in Soweto on depression symptoms is marginally significant (*p* = 0.095). The respective predicted CES-D-10 scores for each group are as provided: Less risk = 5.04, Same risk = 6.25, More risk = 8.17.

Finally, we ran an interaction term (Model 11) between perceived COVID-19 risk and childhood trauma to examine whether early stress altered the association between perceived COVID-19 risk and depression, as we hypothesized that greater childhood trauma would exacerbate the associations between depressive symptoms of COVID-19 risk. Model 11 shows evidence for a marginally significant crossover interaction between childhood trauma and perceived risk (*F*[1, 209] = 3.80, *p* = 0.0617) and accounts for 23% of the variance in depressive symptoms. Figure 2 demonstrates that the depressive impacts of heightened perceived COVID-19 risk were greater among individuals who reported worse histories of childhood trauma, yet negligible in the low risk group. The effects of psychiatric risk, COVID-19 knowledge, and quality of life on CES-D scores were consistent. Additional analyses show that COVID-19 knowledge and perceived COVID-19 risk were inversely and significantly related regardless of educational status, childhood trauma, psychiatric risk, coping ability, quality of life, and demographic factors (β = -0.047, *p* = 0.043, 95% CI [-0.09, -0.003]). An identical logistic regression model showed that the interaction between childhood trauma and perceived risk was not significant, likely due to the non-linearity assumed in logistic models^13^.

**Figure 2.**
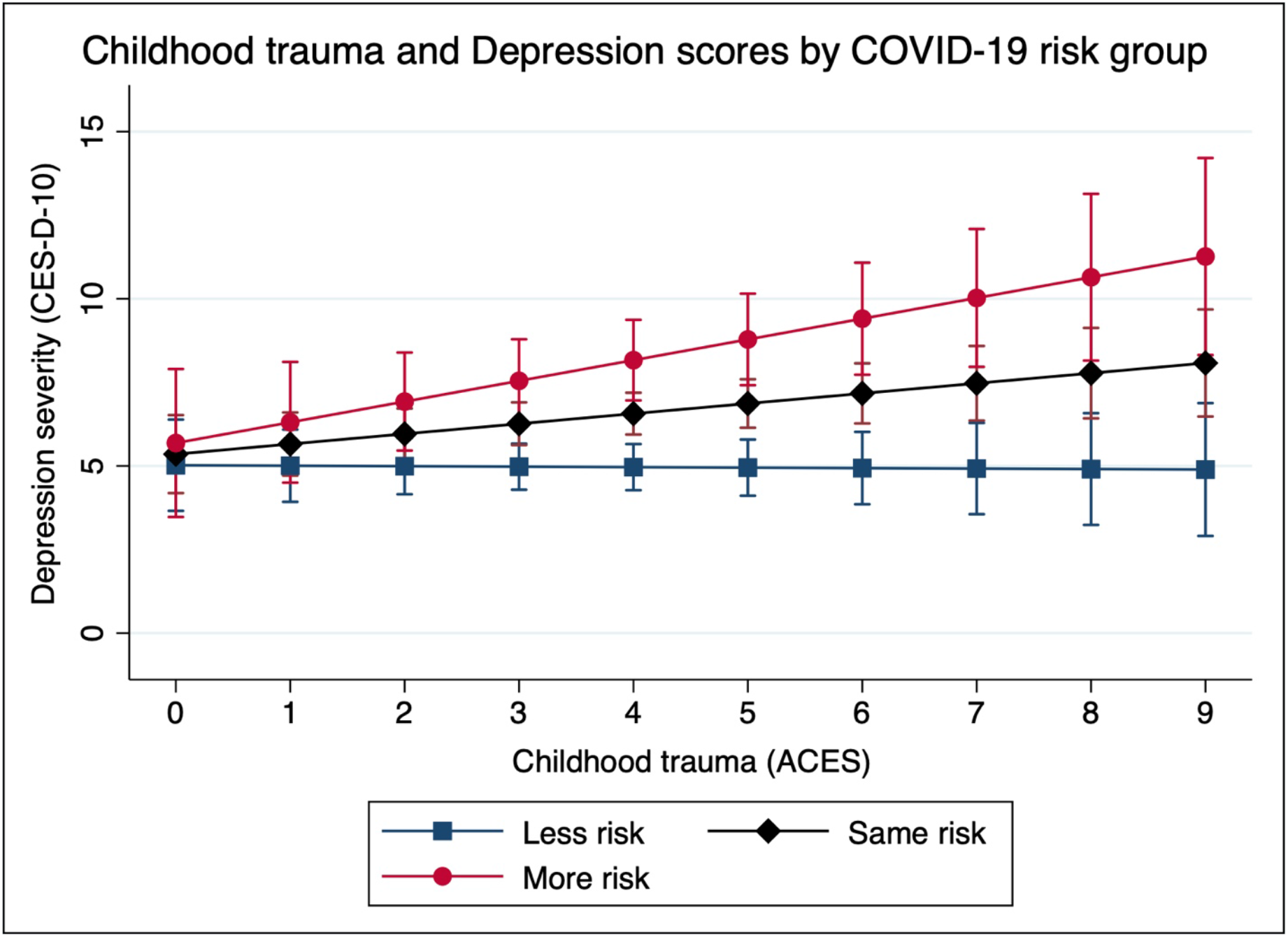
Childhood trauma (ACES) and Depression scores (CESD) by COVID-19 risk group. *Note:* Greater childhood trauma (ACES) potentiates the positive relationship between greater perceived COVID-19 risk and the severity of depressive symptomatology. The effect of the interaction between childhood trauma and perceived COVID-19 risk on depression is marginally significant (*F*[1, 206] = 3.53, *p* = 0.0617).

## DISCUSSION

The South African lockdown effectively prevented possible COVID-19 infections and associated morbidity and mortality risk: epidemiological models suggest the lockdown resulted in a 40-60% reduction in transmission relative to baseline after one month^1^. The mental health consequences caused by the rapid and dramatic societal changes from the lockdown, however, cannot be overlooked in a country with considerable psychiatric morbidity, limited mental healthcare infrastructure, and high rates of poverty and unemployment. Our findings highlight that the depressive impacts of greater perceived COVID-19 infection risk were more severe among individuals who reported worse histories of childhood trauma. Additionally, our study emphasizes the prioritization and provision of accessible mental health resources for resource-limited communities in Soweto and across South Africa.

We discuss three main findings from our analyses. Few cases had been detected in Soweto during this first month of lockdown, although a portion of our sample perceived their risk as high and expressed deep anxiety and fear over personal and family well-being. Our first major finding is that people perceive their own risk for COVID-19 infection differently relative to others in their community. These risk perceptions were unaffected by age, gender, socioeconomic status, educational attainment, household density, coping ability, and whether people had heard of coronavirus and had been tested. Our qualitative data suggest that inability to properly social distance and quarantine, potent psychosocial stress, preexisting health conditions, unemployment, and food insecurity exacerbate existing individual and household strains among already vulnerable families, placing them at greater risk of disease susceptibility. Greater knowledge about the prevention and transmissibility of coronavirus was associated with lower perceived risk of COVID-19 infection. This result emphasizes importance of public health awareness campaigns and effective messaging around prevention strategies as well as the numerous impacts of preexisting structural vulnerabilities on differential perceptions of COVID-19 infection risk. This finding recapitulates the lessons learned from the HIV/TB epidemics in South Africa, that fundamental causes of infectious disease must be systematically prioritized across the continuum of public health responses, from emergency response, treatment, education, to prevention.

Second, while most did not think that COVID-19 affected their mental health, we found a variety of stressors that caused deep worry, anxiety, and rumination (“thinking too much”) in approximately 20% of adults. These constant foci during the lockdown were driven and exacerbated by the inability to care for themselves and their families, crippling financial concerns, personal vulnerability due to illness, the invisible nature of COVID-19 transmission, and a lack of awareness on COVID-19. It is unsurprising that the same factors that motivated concerns over mental distress also impacted one’s perception of their own risk of COVID-19, which highlights the coupling of perceived disease risk and depressives symptoms in our sample. The discordance between the highly prevalent perception in our interviews that COVID-19 did not impact mental health and the strong relationship between perceived risk and depressive symptoms, however, raises concern that participants may not be aware of the potential threats to their mental health during COVID-19.

Third, our main finding shows that higher self-perceived risk of COVID-19 infection is associated with greater depressive symptoms in adults with more severe histories of childhood trauma during the first six weeks of lockdown. This association remained after controlling for recent psychiatric risk, quality of life, COVID-19 knowledge, coping ability, and demographic factors. While the cross-sectional nature of our measures of perceived COVID-19 risk and depressive symptoms limits our ability to determine casual pathways that may precipitate these trends, the long-term impacts of early life trauma on adult risk perception and mental health are notable (Figure 2).

We propose two possible pathways that may explain the stronger depressive effects of heightened perceived COVID-19 risk, and vice versa, in relation to childhood trauma. First, increased severity of childhood trauma may cause durable increases in psychological and physiological stress reactivity into adulthood and increase one’s risk of developing MDD^14,15^. A growing literature has documented that greater exposure to childhood trauma can alter the development of stress physiological mechanisms, such as hypothalamic-pituitary-adrenal axis regulation, the immune system, and brain function, and leave lasting changes that extend across the lifecourse. For instance, individuals with histories of childhood trauma and chronic stress across development exhibit greater risk for increased psychological stress reactivity^16^, heightened cortisol reactivity^17^, elevated inflammatory profiles^18^ in response to future stressors. Recent evidence has reported the long-term impacts of childhood maltreatment on brain regions, such as the amygdala^19^ and hippocampus^20^, that regulate the perceptions of threat appraisal and emotions (e.g. fear, sadness) and are involved in the pathogenesis of MDD^21^ and that epigenetic modification of genetic loci may underlie these processes^22^. These early life stress-linked alterations in stress physiology may subsequently predispose individuals to developing a suite of psychopathologies, including depression.

Conversely, greater past childhood trauma may increase the severity of adult depressive symptoms or MDD and increase emotional and biological sensitization to future stressors and adverse conditions. Childhood trauma is a well-known risk factor that influences the severity and duration of MDD and other psychopathologies^23,24^. Additionally, key symptomatic behaviors of MDD, such as persistent feelings of victimization, learned hopelessness and helplessness, and negative appraisal^25,26^ likely motivate the elevated self-perception of COVID-19 infection risk. Adults with histories of childhood trauma and greater MDD severity, particularly among melancholic MDD patients, have developed increased psychological^26^ and neuroendocrine^17^ sensitization to future stressors. Previous research on perceived AIDS risk found that adult women with greater severity of childhood trauma reported increased perceptions of contracting HIV^28^. Thus, elevated perceived COVID-19 infection risk may arise as a function of the depressive effects from childhood trauma. Future longitudinal research is needed to determine the precise mechanisms by which childhood trauma, perceived COVID infection risk, and depressive symptoms are related.

While we have examined the effects of a variety of individual-level factors across the lifecourse on perceived infection risk and depressive symptoms, the social and historical contexts from which many of these social and psychological factors arise are largely responsible for the current state of psychiatric morbidity and vulnerability to infection in Soweto today. Nearly all participants were born during the oppressive apartheid regime or shortly after its violent dissolution, which is when all reported childhood traumas took place. Though children were not always exposed to the everyday adversities and extreme traumas of racial segregationist cultures and policies, the distributive impacts of racialized and classed violence among families often times translated to poor housing quality, food insecurity, family violence, and child abuse^29,30^. The psychological, economic, and structural legacies of apartheid violence manifest in the present moment where the intergenerational trauma of apartheid may persist and sustain racial and class disparities in mental illness, socioeconomic opportunity, and infectious disease risk. We offer this history to contextualize our findings and emphasize the importance of prioritizing accessible mental health and infectious disease prevention services countrywide.

## CONCLUSION

To the best of our knowledge, this is the first investigation of the mental health impacts of COVID-19 experiences during the 2020 coronavirus pandemic and national lockdown in South Africa. We report that the relationship between increased depressive symptoms and greater perceived COVID-19 infection risk was more severe among adults who reported worse histories of childhood trauma. Adults were two times more likely to experience significant depressive symptoms for every one unit increase in perceived COVID-19 risk. Greater knowledge of COVID-19 prevention and transmission was associated with lower perceived risk of depression but higher depressive symptoms. While a large majority of participants reported that experiences of the COVID-19 pandemic did not affect their mental health (or “mind”), 10-20% of participants reported potent experiences of anxiety, fear, and “thinking too much” as a result of the pandemic. Our results highlight the compounding effects of past traumatic histories and recent stress exposures on exacerbating the severity of depressive symptoms among adults living in an urban South African context.

## Data Availability

Data are available upon request.

## Funding

The research was funded by a grant (1R21TW010789-01A1) to EM from the Fogarty International Center at the US National Institutes of Health. AWK is supported by the National Science Foundation Graduate Research Fellowship and the Fogarty International Center and National Institute of Mental Health, of the National Institutes of Health under Award Number D43 TW010543. The content is solely the responsibility of the authors and does not necessarily represent the official views of the National Institutes of Health.

### Acknowledgements

We are greatly indebted to the participants, their families, and our research assistants as this study would not have existed without them. Specifically, we would like to thank Lindile Cele, Sbusiso Kunene, Gladys Morsi, Sharlotte Sihlangu, and Jackson Mabasa for their hard work calling our study participants during the early days of lockdown. We are also extremely grateful for the front-line and community health workers who are working endlessly to keep Soweto and the rest of South Africa healthy and safe.

## Author contributions

AWK conceptualized the question, study design, and analysis, oversaw data collection, conducted quantitative data analysis, and drafted the manuscript. TN assisted with data cleaning and analysis and reviewed the paper. EM acquired funding for the parent study, designed the study protocol, oversaw and conducted qualitative data analysis, and reviewed the manuscript.

## Footnotes

* The initial version of this question queried how COVID-19 affects mental health, but “mental health” was not a well-understood phrase. Rather, we found that using the term “mind” indexed identical meanings to mental health and was more intelligible in English and when translated to isiZulu, isiXhosa, and Sesotho.

